# Direct modelling from GPS data reveals daily-activity-dependency of effective reproduction number in COVID-19 pandemic

**DOI:** 10.1101/2022.09.27.22280400

**Authors:** Jun’ichi Ozaki, Yohei Shida, Hideki Takayasu, Misako Takayasu

**Author notes:** Electronic Address.

## Abstract

During the COVID-19 pandemic, governments faced difficulties in implementing mobility restriction measures, as no clear quantitative relationship between human mobility and infection spread in large cities is known. We developed a model that enables quantitative estimations of the infection risk for individual places and activities by using smartphone GPS data for the Tokyo metropolitan area. The effective reproduction number is directly calculated from the number of infectious social contacts defined by the square of the population density at each location. The difference in the infection rate of daily activities is considered, where the ‘stay-out’ activity, staying at someplace neither home nor workplace, is more than 28 times larger than other activities. Also, the contribution to the infection strongly depends on location. We imply that the effective reproduction number is sufficiently suppressed if the highest-risk locations or activities are restricted. We also discuss the effects of the Delta variant and vaccination.

## Introduction

Since the beginning of the COVID-19 pandemic in 2019, there have been 452 million confirmed cases and over six million deaths globally as of 12 March 2022, posing serious healthcare challenges^1–3^. Most governments have struggled to control this disease and simultaneously minimize its damage to daily and economic activities owing to limited time and resources^4–8^. Along with vaccinations, nonpharmaceutical interventions are considered essential for managing this disease^9–13^. For example, the governments around the Tokyo metropolitan area in Japan declared states of emergency (SoEs) that limited daily and economic activities including schools, department stores, cinemas, restaurants, bars, and travel to reduce human mobility in public spaces^5,9^. Consequently, the number of social contacts and, thereby, the effective reproduction number decreased. Governments require reliable quantitative estimations of the effect of such policies on the pandemic, that is, a predictable model, for making informed decisions.

Studies have estimated the effectiveness of lockdowns or non-compulsory measures such as SoEs on reducing human mobility^9–13^. However, the effect of reduced human mobility on the pandemic remains unclear, as it is insufficient to investigate the number of social contacts alone. Infectious diseases, including the COVID-19 pandemic, have been studied using global positioning system (GPS) data^14–23^. However, with such diseases, the types of social contacts are considered more critical. Specifically, the infection rate is known to strongly depend on whether social contacts are wearing masks or talking and on the ventilation state of the rooms they are in^24–27^. The point is that the number of social contacts through each daily activity type should be investigated.

In this study, we propose a model to predict the effective reproduction number of COVID-19 based on daily human activities inferred from smartphone GPS data. From these data, first, we categorize citizens’ daily activity patterns into four types and estimate the population density for each activity, location, and time in the Tokyo metropolitan area. We also calculate the number of social contacts for each activity by assuming it to be proportional to the square sum of the population density. Second, we propose an activity-dependent infection model based on the susceptible-infectious-removed (SIR) model. This model seems to have a lower resolution than that of other compartmental models (e.g., susceptible-exposed-infectious-removed (SEIR) model). However, it is suitable for direct formulation using GPS data. The effective reproduction number is a linear combination of the number of social contacts, and the coefficients are the infection rate per contact. We determine the parameters to fit the empirical data before the spread of the Delta variant and verify that the model is sufficient to predict the effective reproduction number. Then, we compare the parameters to observe the activity that has the highest infection risk. We also calculate the effective reproduction number for each daily activity, location, and individual. The model prediction is valid for up to around two weeks after the human mobility data are obtained. Third, we investigate effects other than those of human mobility. We show that the domination of the Delta variant is described by only two parameters: starting date and ratio of effective reproduction number to that of existing variants. Furthermore, we demonstrate the effectiveness of vaccination through its high prevention ratio of infection in a metropolis.

## Methods

### Epidemiological data

COVID-19 epidemiological data were provided by the Ministry of Health, Labour, and Welfare in Japan^28^. The data consist of the number of new infection cases and severe cases from Feb. 2020 and May 2020, respectively, to Oct. 2021. We took a 7-day moving average to remove the periodicity of epidemiological data in a week. The moving average was taken from 6 days prior to the target date. Vaccination statistics were obtained from the Prime Minister of Japan and His Cabinet^29^.

### GPS data

Our GPS data from 1 Jan. 2020 to 30 Sep. 2021 were purchased from Agoop Corp, Japan^30^, which collected the information of smartphones’ location only if the users downloaded some smartphone applications provided by Agoop and gave permission and their consent to the terms of use for the applications following the Privacy Policy of Agoop Corp^31^. The example and description of the data are given in Supplementary note 2. The data were fully anonymized. Namely, all user IDs were renewed every midnight, so any tracking of individual persons across the days is impossible. The median accuracy of the latitude/longitude data was 10m. However, the precise home location was masked by limiting the data resolution around their home for privacy protection in the following way. If a user is located in a 1km-square where their home is located, the location data was replaced by that of the centre of the square; also, the size of the square extends to 10km-square in low population areas. This square size is used only for the data provider to mask the users’ homes in very low population areas. As for users’ workplaces, the city’s name is given in the resolution of municipalities. The GPS data has a bias in generations: the Ministry of Internal Affairs and Communications, Japan, reported that the ratio of the smartphone users is concentrated between 13-59 ages in 2019^32^. A bias in the home location is discussed in Supplementary note 3. We confirmed that the population observed in the GPS data is similar to the actual population in the census data, although the GPS data has a bias to concentrate on the high-density population areas more than the census data. The anonymized GPS data is commercially available from Agoop Corp., which complies with the Japanese Personal Information Protection Act. Human Subjects Research Ethics Review Committee of Tokyo Institute of Technology authorized that no ethical review is needed in this case.

We pre-processed the original data to obtain 15 min interval data using linear interpolation, discarding data that cannot be interpolated sufficiently. The timestamps were every 15 min from 05:00 to 24:00. Data for midnight were discarded because most people stay in bed and do not spread infection. The home/work city data were converted into the home/work 1 km-square data. A home square is a 1 km-square where the user stays at 05:00 in the home city, and a work square is where the user visits for at least 5 h in a day in the work city. Data for iOS smartphones were discarded because they are not gathered if the user is not moving due to the iOS specification, making staying at home undetectable. The converted data cover approximately 0.4 million users in Japan. We investigated the population corresponding to a GPS point to renormalize the GPS data to the actual population distribution. We counted the users at 05:00 for each home prefecture defined by their home city’s prefecture for each day. The effective population of one GPS data point in a prefecture was calculated as the ratio of the actual population in the prefecture in Oct. 2019^33^ to the number of users counted. At the beginning of 2020, the typical values ranged from 250 in metropolitan areas to 500 in the countryside.

## Results

### Epidemiological data

First, we analysed COVID-19 epidemiological data in the Tokyo metropolitan area (Tokyo, Kanagawa, Saitama, Chiba, Ibaraki, Gumma, Tochigi, and Yamanashi prefectures) from 1 Feb. 2020 to 31 Oct. 2021^28^. *t* denotes the day count from the beginning of 2020 (e.g., *t* = 1 means 1 Jan. 2020) and *I*^new^(*t*), the number of new COVID-19 infection cases on day *t*. The effective reproduction number 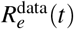 was estimated as^34^

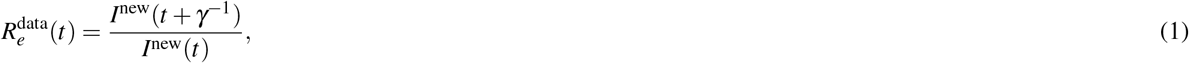

where *γ*^−1^ = 5 is the mean generation time^35^. Fig. 1(a) shows a plot of the effective reproduction number. Before 30 Mar. 2020, the data is unreliable because the number of PCR tests per day is under 10,000 in this period, and the confirmation of infection is not considered comprehensive. In this span, the effective reproduction number was estimated using an average of 40 days. Assuming the effective reproduction number is constant as 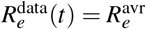, Eq. (1) results in 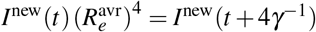. Here we sum the number of newly confirmed cases over the former 20 days and the latter 20 days of the 40 days. Then, we 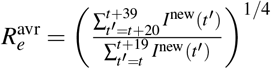, where we did not take a 7-day moving average for the calculation of *I* ^new^ (*t*). That is, the average effective reproduction number of the 40 days is determined by the ratio of the number in the latter 20 days to that in the former 20 days to the power of 1/4. The difference from the estimation by the Cori method^36^ is shown in Supplementary note 4. The result of the method in this study is consistent with that of the Cori method. Similarly, we plotted the effective reproduction number limited in severe cases, which is defined using only the number of new severe cases instead of the number of all infections. The correlation between the effective reproduction numbers for all cases and severe cases is maximized if that for severe cases is regarded to be delayed by 12 days. Both are consistent with each other after *t* = 200. The 1st to 4th SoEs in the Tokyo metropolitan area^5^ are also shown. During each SoE, the effective reproduction number started to decrease after around two weeks. The Delta variant was first detected in Japan on 18 May 2021 (i.e., *t* = 504)^37^.

**Figure 1.**
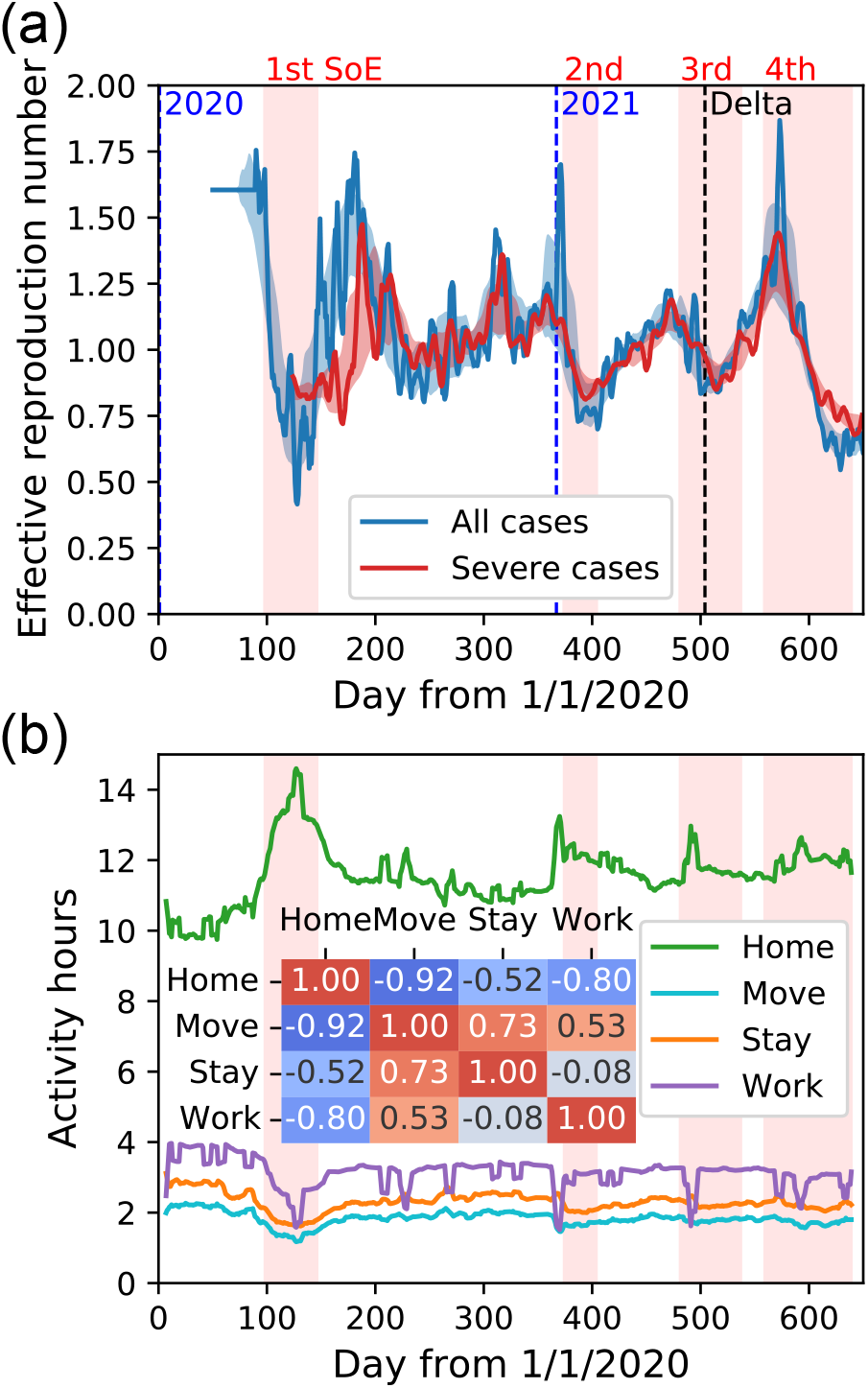
(a) Effective reproduction number of all infections and severe cases in the Tokyo metropolitan area. The SoEs and beginnings of years are also shown. The first case of the Delta variant in Japan is represented as ‘Delta’. Severe cases are plotted, taking into account the 12-day delay. The light-colored range is estimated as follows. The centre of the range is the 31-day moving average of the original time series, and the range width is the 31-day moving average of the absolute difference between the centre and original time series. Before 30 Mar. 2020, the effective reproduction number is estimated using an average of 40 days because the confirmation of infection is not considered comprehensive. (b) Time series of mean duration per day of each activity in the Tokyo metropolitan area. The 7-day moving average is taken. The 1st–4th SoEs are also shown. The Pearson’s correlation among the mean duration of the four activities in the span 200 ≤ *t <* 500 is calculated.

### Human mobility and activity patterns

We analysed empirical human mobility data in Japan, especially in the Tokyo metropolitan area, from 1 Jan. 2020 to 30 Sep. 2021^30^. We calculated the population density at each time and location to estimate the number of infection routes. The infection probability of each infection route depends on how people contact each other and, consequently, on the activity. Therefore, we categorized the GPS user activity into four states, staying home (‘home’), moving (‘move’), staying out (‘stay’), and working (‘work’), to distinguish the activity-dependent infection probability with a time resolution of 15 min. The definition is as follows: (a) home is the state when the user is within the 1 km-square corresponding to their home (home square), (b) work is the state when the user is within the 1 km-square corresponding to their workplace (work square), (c) move is the state in which the user is within a 1 km-square that is different from the one in which they were 15 min ago and that does not correspond to either their home or workplace, and (d) stay-out is any other situation, e.g., in restaurants, department stores, or stadiums away from their home. Here, the home and work locations (home/work square) are estimated from the home/work city data in the GPS dataset, as explained in the *Methods* section. For an example of the categorization, if a user is at home/work 1 km-square, then the activity type is home/work (home is preferred if both are the same). If a user moves from another 1 km-square, the activity is move unless the following location is not home/work square. Even if the user stays for over 5 hours, the activity type is not work in a location other than the work city. Note that home activity is not limited to staying at home literally. It also includes activities within a 1 km-square around the home location, such as shopping and having a meal at a restaurant.

Fig. 1(b) shows a plot of the time series of the mean duration per day of the four activities, where we take the 7-day moving average for avoiding the periodicity related to weekdays. The total time per day is 19 h because the data point is from 05:00 to 24:00. During the pandemic, roughly speaking on average, users were at home for 12 h (not including midnight), at work for 3 h, moved for 2 h, and stayed out for 2 h. Note that this is averaged over all days including holidays for all users. We also calculated the correlation among the mean durations of the four states in the span 200 ≤ *t <* 500. This span was used later for model parameter fitting. Move was strongly correlated with work and stay.

### Model

We derived an infection model based on the classical SIR model using the GPS data for the four states with several assumptions. The classical SIR model is given by the following set of equations:

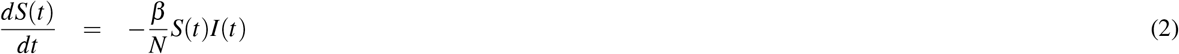

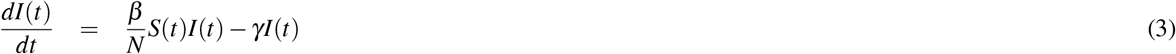

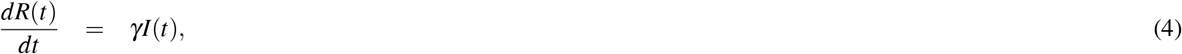

where *t* is the time; *S*(*t*), the number of susceptible people; *I*(*t*), the number of infectious people; *R*(*t*), the number of removed people (either recovered, quarantined, or dead); and *N* = *S*(*t*) + *I*(*t*) + *R*(*t*), the total number. The parameter *β* represents the strength of infection spread, and the parameter *γ* determines the timescale from infected state to recovered or quarantined state. The reciprocal of *γ* is the mean generation time^35^: *γ*^−1^ = 5. We adopted the following four assumptions:

(A1) the number of removed people is much smaller than the number of susceptible people,
(A2) no nonlocal infections occur between different spaces or time periods,
(A3) infectious people are distributed uniformly in the target area (i.e., Tokyo metropolitan area), and
(A4) no infections occur between different activities.

Assumption (A1) comes from the low cumulative number of COVID-19 cases in Japan (below 2% on 30 Oct. 2021)^28^. Then, the number of susceptible people is assumed to be constant, that is, the whole population of Japan. Assumption (A2) means that we discard indirect infections such as droplet infection with long-distance or contact infection after a long time^38^. Nonetheless, indirect infections are effectively included if the population in the target area does not change drastically. Assumption (A3) is used for simplifying the model. As our GPS data does not contain users’ privacy including the information of infection, we simply assume that infected people distribute uniformly. We discard the effect from infected people who went outside or inside of the Tokyo metropolitan area. Assumption (A4) is the intuition that social contacts between different activities are fewer than those within the same activity. In the Tokyo metropolitan area (and possibly most other metropolitan cities), the office space and restaurants are separated even though they are on the same 1km-square. Working people in office towns are considered to have a comparably small interaction with the stay-out people because most of them are working in their offices. In addition, the workers contacting with stay-out people (e.g., waiters contacting customers in restaurants) are strongly recommended to wear masks^5^ and considered to be less infectious.

By applying the first assumption (A1), we only need to consider the second equation of the SIR model because infection causes a negligible change in *S*(*t*). In this case, the equation becomes

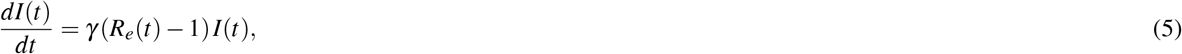

where 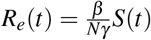 is an effective reproduction number. We discretize the equation by setting *dt* = *γ*^−1^ as

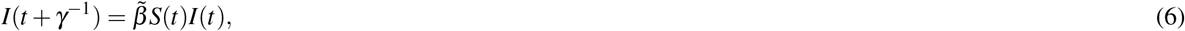

where 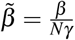. This discretization is needed for fitting the empirical data.

Next, we adopt the second assumption (A2) of local infections. We suppose that all people in the target area have equal possibility of close contact with each other. In this case, the time evolution of *I*(*t*) is described by the product of *I*(*t*) and *S*(*t*) because the number of infection routes or social contacts is proportional to *I*(*t*)*S*(*t*). In this sense, the classical SIR model is exact only if the population density is uniform. However, this is not the case in real situations. Therefore, we divide the area into squares (grids) in which social contacts are equally possible among all people in a square. The size of the squares is set to 1 km×1 km. The case of 100 m×100 m^9^ is discussed in Supplementary note 1. Thus, we obtain

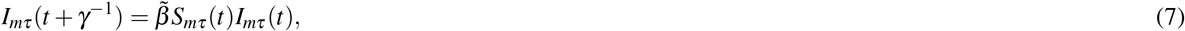

where *m* is the square label, and *τ* is the time label in the considered area *A* and date *t*. The area *A* is the Tokyo metropolitan area, and it contains 36,898 1 km-squares. The time label *τ* moves in 15 min intervals around the current date *t* for five days, from *t* − 2 to *t* + 2, according to the discretization unit *γ*^−1^ = 5. The parameter 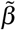 takes a different value from the earlier equations because the space size and period are different: 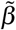 is multiplied by (number of 1 km-squares)/(number of time steps) if the density is uniform in space and time.

The third assumption (A3) gives the following relation:

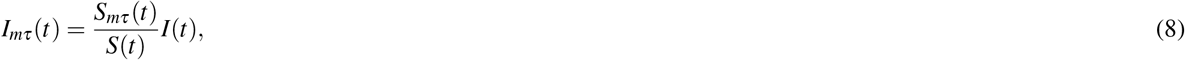

where *S*(*t*) = ∑_*m*∈*A*_ *S*_*mτ*_ (*t*), *I*(*t*) = ∑_*m*∈*A*_ *I*_*mτ*_ (*t*), and *A* is the target area. Therefore, the effective reproduction number is given as

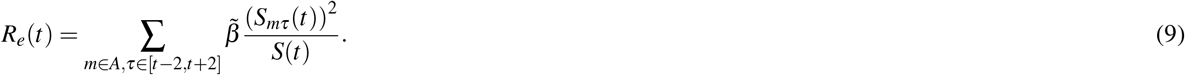

Finally, assumption (A4) is applied. We divide the population density *S*_*mτ*_ (*t*) into that of the activities *S*_*amτ*_ (*t*) and introduce activity-dependent parameters *β*_*a*_. The equation is

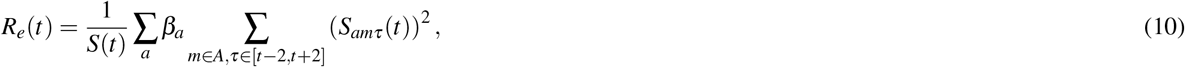

where the time label *τ* moves from *t* − 2 to *t* + 2, and the activity label *a* takes the values ‘home’, ‘move’, ‘stay’, and ‘work’. The coefficient *β*_*a*_ is supposed to be constant in the timespan considered. For simplicity, we approximate the sum as follows:

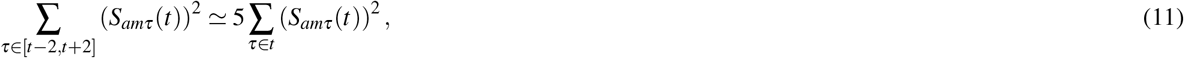

for the equation to have values only on date *t*, where the time label *τ* moves only on date *t* in the right-hand side. Thus, the effective reproduction number is a linear combination of the human mobility time series,

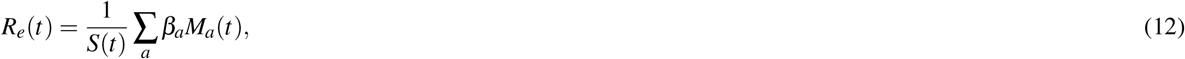

by introducing *M*_*a*_(*t*) = *γ*^−1^ ∑_*m*∈*A,τ*∈*t*_ (*S*_*amτ*_ (*t*))^2^, the population moment of activity *a* on date *t*. We emphasize that the effective reproduction number is simply determined only by the human mobility on the same day. The parameters *β*_*a*_ are interpreted as the infection rate during activity *a* per infection route and 15 min period.

We determine the model parameters *β*_*a*_ to fit the effective reproduction number; however, the observed effective reproduction number is based on the report date and not the infection date. We have to consider a typical time delay from infection to report Δ*T* to deal with it. In light of this effect, Eq. (12) is modified as follows:

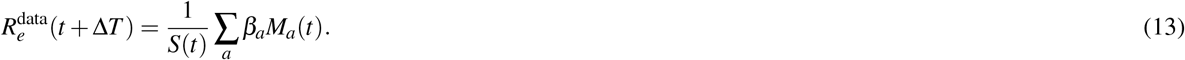

Here, we use the 7-day moving average of *M*_*a*_(*t*) to remove the periodicity of weekdays as well as the definition of the empirical effective reproduction number. We estimate the time delay from infection to report Δ*T* in the period 200 ≤ *t <* 500. Figure 2 shows the correlation between the effective reproduction number and the population moment of each activity delayed by Δ*T*. The peak at Δ*T* ∼ 14 is the delay of the infection reports, whereas the trough at Δ*T* ∼ −60 means that human mobility is decreased after infection spread. The optimal value Δ*T* = 14 is derived by minimizing a fitting loss to the effective reproduction number by the population moments as a function of Δ*T*, where the fitting loss is the squared distance in the log-10 space, to prevent the estimation from being dominated only by the significant value of the effective reproduction number. This value is consistent with that reported in a previous study^39,40^.

**Figure 2.**
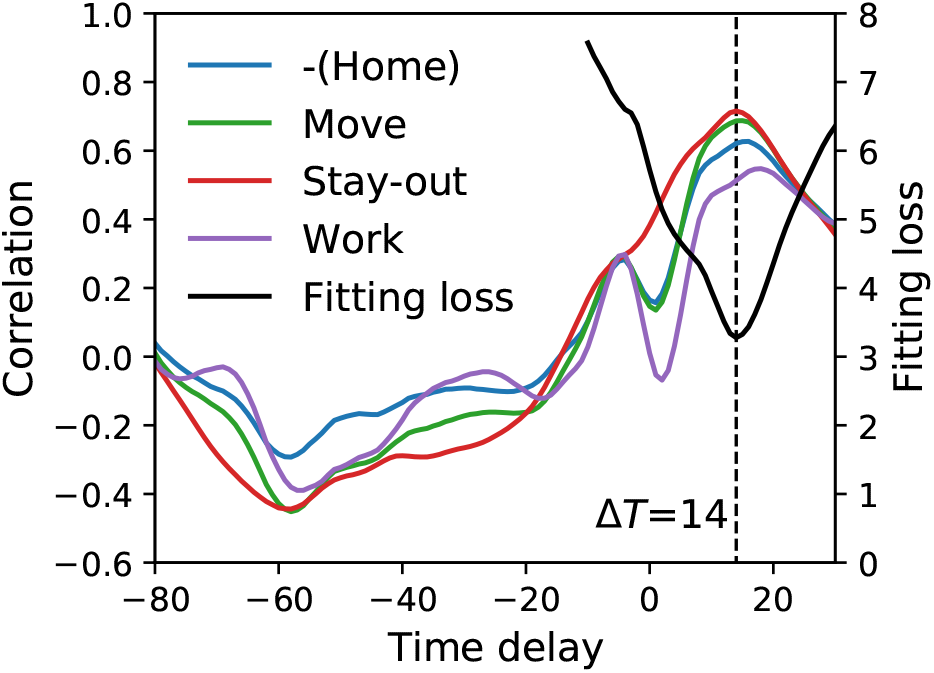
Correlation between the effective reproduction number and the population moments time series delayed by Δ*T*, and fitting loss to the effective reproduction number by the population moments as a function of the delay Δ*T* in the period 200 ≤ *t <* 500. The fitting loss is calculated as the residual of the square sum in the log space. For the correlation, the peak and trough are observed at around Δ*T* = 14 and Δ*T* = −60, respectively. For the fitting loss, it is minimized at Δ*T* = 14.

The remaining problem is the multicollinearity of the move population moment; the correlation of the population moments is 0.89 between move and stay-out and 0.90 between move and work. To address this, we assume that the coefficient *β*_*a*_ for move and work is the same because the situations in these two activities are similar: people wear masks in trains (move) and offices (work) but not always in restaurants (stay-out).

We fit the model parameters to the data for all cases in the period 200 ≤ *t <* 500. We do not use data for *t <* 200 because the social situation drastically changed during the 1st SoE (e.g., mask distribution in Japan^41^), and the data are not stationary. Furthermore, the effective reproduction numbers for all cases and severe cases are different: epidemiological data are not reliable in that span. The fitting is performed under the log space to prevent the estimation from being dominated only by the significant value of *R*_*e*_(*t*). As a result, the parameters are *β*_home_ = (1.2±0.1)×10^−7^, *β*_move_ = *β*_work_ = (6.2±2.8)×10^−8^, and *β*_stay_ = (3.4±0.2)×10^−6^, where the unit is per 15 min. Figure 3 shows that the model explains the data in the period; however, they are different before *t* = 200 and after *t* = 500. Before *t* = 200, the effective reproduction number for severe cases is more consistent with the model result. After *t* = 500, the effects of the Delta variant and the vaccination result in differences^29,37^. Thus, the classical SIR model is suitable for direct GPS data modelling. Despite its simplicity, we observe that the model quantitatively explains the COVID-19 epidemic.

**Figure 3.**
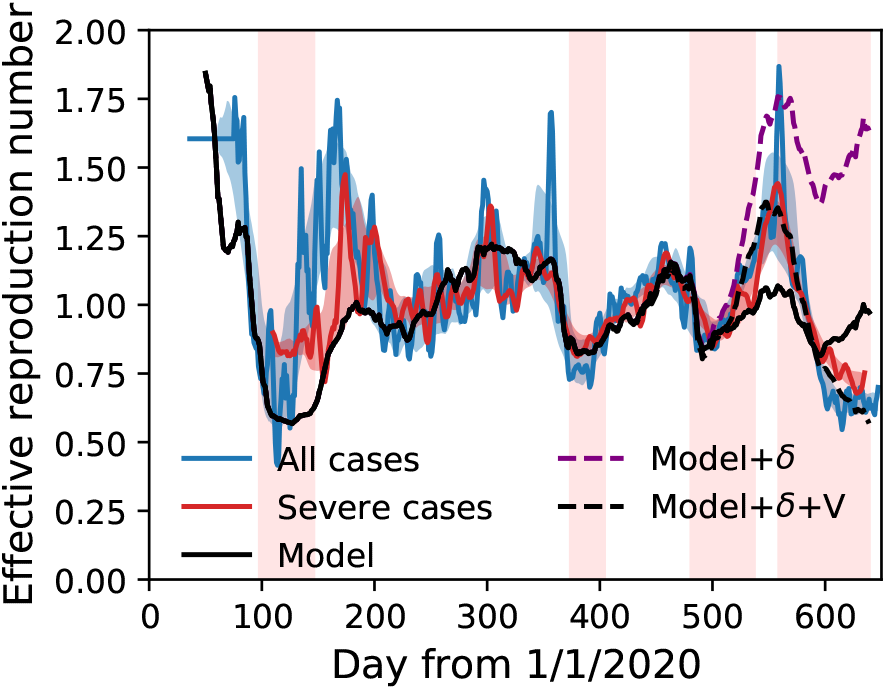
Effective reproduction number of all infections and severe cases in Tokyo metropolitan area compared to the model result. The effective reproduction number is plotted with a delay of 14 and 26 days for all and severe cases, respectively. Parameter fitting is done in the span 200 ≤ *t <* 500. ‘Model’ indicates the prediction from human mobility alone, ‘Model+*δ* ‘ includes the effect of the Delta variant, and ‘Model+*δ* +V’ additionally includes the effect of the vaccination.

Next, we change the threshold concerning the activity duration to investigate the sensitivity of the fitting parameters. As explained in the *Methods* section, a work 1 km-square has been detected by the user’s dwelling for at least 5 hours in their work city. Here, we check the cases of 4 hours and 6 hours minimum dwelling time. The parameters for the 4 hours, 5 hours, and 6 hours cases are obtained in Table 1. The estimated values of *β*_home_ and *β*_stay_ are considered to be unchanged within the margin of the standard deviation, while the values of *β*_move_ and *β*_work_ show a tendency to take smaller values for longer minimum dwelling time. This difference is because the infection rate in move and work is much lower than the other parameters and has a more significant fluctuation relatively. If we decrease the minimum dwelling time, the work hour tends to be counted more. For example, the spent time percentages (move, stay-out, work) in public are (25%, 32%, 42%) for the 4-hour case, (26%, 34%, 40%) for the 5-hour case, and (26%, 36%, 38%) for the 6-hour case, on the average of a week starting from 9 to 15 January 2020, in which COVID-19 did not affect the human mobility.

**Table 1.**
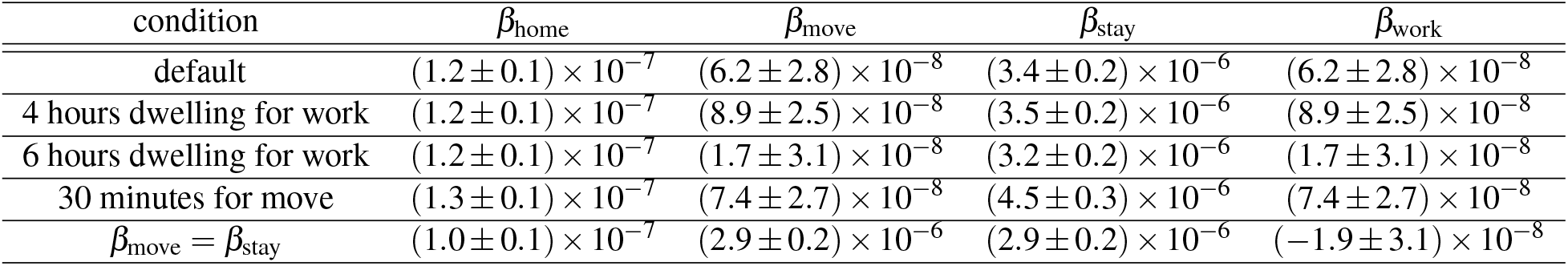
Sensitivity analysis of the fitting parameters of the effective reproduction number. The default minimum dwelling time for detecting work square is 5 hours. The cases of 4 hours and 6 hours are indicated by ‘4 hours dwelling for work’ and ‘6 hours dwelling for work’. ‘30 minutes for move’ represents the case that the minimum moving time is 30 minutes. ‘β_move_ = β_stay_’ is the case that the parameters are jointly estimated for stay-out and move, instead of for work and move.

| condition | $\beta_{\text{home}}$ | $\beta_{\text{move}}$ | $\beta_{\text{stay}}$ | $\beta_{\text{work}}$ |
| --- | --- | --- | --- | --- |
| default | $(1.2 \pm 0.1) \times 10^{-7}$ | $(6.2 \pm 2.8) \times 10^{-8}$ | $(3.4 \pm 0.2) \times 10^{-6}$ | $(6.2 \pm 2.8) \times 10^{-8}$ |
| 4 hours dwelling for work | $(1.2 \pm 0.1) \times 10^{-7}$ | $(8.9 \pm 2.5) \times 10^{-8}$ | $(3.5 \pm 0.2) \times 10^{-6}$ | $(8.9 \pm 2.5) \times 10^{-8}$ |
| 6 hours dwelling for work | $(1.2 \pm 0.1) \times 10^{-7}$ | $(1.7 \pm 3.1) \times 10^{-8}$ | $(3.2 \pm 0.2) \times 10^{-6}$ | $(1.7 \pm 3.1) \times 10^{-8}$ |
| 30 minutes for move | $(1.3 \pm 0.1) \times 10^{-7}$ | $(7.4 \pm 2.7) \times 10^{-8}$ | $(4.5 \pm 0.3) \times 10^{-6}$ | $(7.4 \pm 2.7) \times 10^{-8}$ |
| $\beta_{\text{move}} = \beta_{\text{stay}}$ | $(1.0 \pm 0.1) \times 10^{-7}$ | $(2.9 \pm 0.2) \times 10^{-6}$ | $(2.9 \pm 0.2) \times 10^{-6}$ | $(-1.9 \pm 3.1) \times 10^{-8}$ |

We also estimate the sensitivity of move activity. For explanation, here we suppose the case that the user is not at home or work. In this supposition, move is the state in which the user is out of a 1 km-square where they were located 15 min ago. This definition means that the minimum moving time is evaluated as 15 minutes. Instead, we consider the interpretation that the minimum moving time is 30 minutes. We append an additional criterion for move that the 1 km-square in the next 15 minutes is different from the present. For instance, if a user is in A at 6:00 and B at 6:15, the state is move both at 6:00 and 6:15 under this criterion, while it is move only at 6:15 under the previous criterion. The result in this case is shown in Table 1. The parameter *β*_stay_ is increased because the effective reproduction number is supported by the decreased stay-out time. The ratio *β*_stay_*/β*_home_ is increased to about 35.

In addition, we check the case with the assumption of the relation, *β*_move_ = *β*_stay_. We have investigated the case of *β*_move_ = *β*_work_ because of the multicollinearity of the population moment of move with that of stay-out and work. If we assume the opposition case *β*_move_ = *β*_stay_ instead, the parameters are changed as in Table 1. As a result, the parameters for home, work and stay decrease, while the parameter for move soars. The ratio *β*_stay_*/β*_home_ = 29 is still close to that in the case of *β*_move_ = *β*_work_.

### Delta variant and vaccination effects

The effects of the Delta variant and vaccination are estimated as follows^42^. Let *r*_*δ*_ = *R*^*δ*^ */R*^0^ be the ratio of the effective reproduction number of the Delta variant *R*^*δ*^ to that of the other existing variants *R*^0^. Furthermore, we assume that the numbers of people infected by the Delta variant *I*^*δ*^ (*t*) and the existing variants *I*^0^(*t*) are the same at *t* = *t*_*δ*_. If both variants increase independently, the ratio is 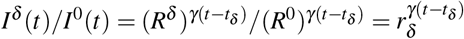. By definition, the effective reproduction number of mixed variants *R*^mix^(*t*) is

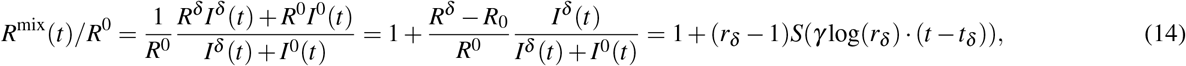

where *S*(*x*) = 1*/*(1 + exp(−*x*)) is the standard sigmoid function. The range of *R*^mix^(*t*) is *R*^0^ to *R*^*δ*^. Therefore, the effect of the Delta variant is introduced by multiplying by the factor 1 + (*r*_*δ*_ −1)*S*(*γ* log(*r*_*δ*_)·(*t* −*t*_*δ*_)). The vaccination model is also multiplied by a factor

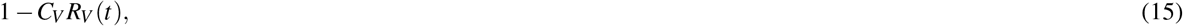

where *C*_*V*_ is the infection prevention ratio of the vaccine^43,44^, and *R*_*V*_ (*t*) is the vaccination ratio in the target area. We approximate the vaccination ratio as *R*_*V*_ (*t*) =(number of vaccinations in Japan up to date *t*)/2(Japanese population) and fit its data by a sigmoid function

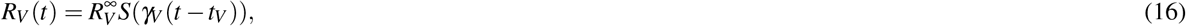

as shown in Fig. 4. The parameters are 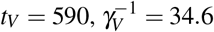, and 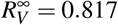. The sigmoid function *S*(*x*) is suitable for fitting the vaccination ratio because it is saturated in the limit *x* → ±∞.

**Figure 4.**
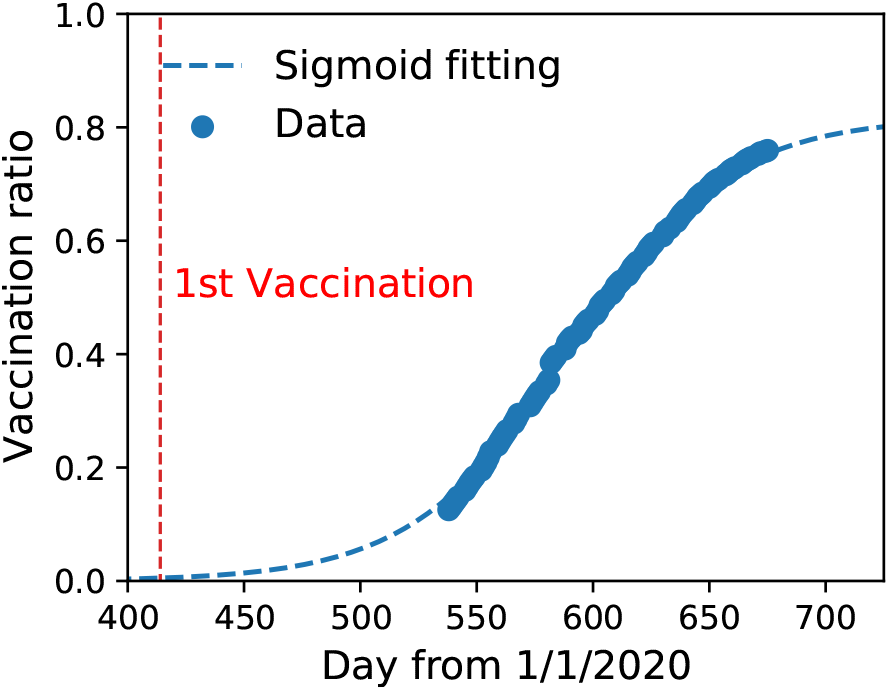
Vaccination ratio of the population of Japan calculated as *R*_*V*_ (*t*) =(number of vaccinations in Japan up to date *t*)/2(Japanese population). The first vaccination day is shown. A sigmoid-like function 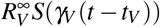 fits the data, and the parameters are 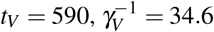, and 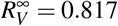.

We determine the other parameters of the Delta variant and the vaccination after *t* = 500 by fitting the epidemiological data as *r*_*δ*_ = 1.68 ± 0.03, *t*_*δ*_ = 530 ± 2, and *C*_*V*_ = 0.99 ± 0.02, where the model equation is finally

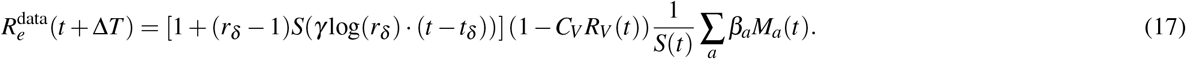

Figure 3 implies that the effects of the Delta variant and the vaccination are fully explained. The values of the parameters, effective reproduction number ratio of the Delta variant, and vaccination’s effectiveness are comparable to those reported previously^42–46^.

### Components of effective reproduction number

The effective reproduction number for each activity, location, and person was investigated. Fig. 5 shows the result for each activity: the sum equals the total effective reproduction number. The stay-out activity dominates the change in the whole effective reproduction number because the parameter for stay-out is 28 and 55 times larger than that for home and move/work, respectively. The effective reproduction number at each location *m* is defined by the partial sum of Eq. (12)

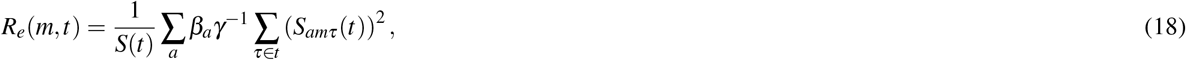

as shown in Fig. 6 (a) two months just before the 1st SoE, (b) during the 1st SoE, (c) two months just before the 2nd SoE, and (d) during the 2nd SoE. The movie of the effective reproduction number map each day is provided as Supplementary movie 1, where a raw value is taken instead of a 7-day moving average to calculate the population moments. The sum over the Tokyo metropolitan area gives the total effective reproduction number. High-risk zones are concentrated in downtown Tokyo. Fig. 7 shows the cumulative distribution. This figure shows that the distribution is almost the same for low-risk regions, and a power law approximates it for high-risk regions whose exponents are (a) -0.93, (b) -1.73, (c) -1.03, and (d) -1.12. Exponents close to -1 imply that the highest-risk regions dominantly affect the total effective reproduction number. In fact, the top-20 highest-risk 1 km-squares in the Tokyo metropolitan area (36, 898km^2^) contribute (a) 40%, (b) 13%, (c) 32%, and (d) 27% of the total effective reproduction number. Consequently, restrictions in the highest-risk downtown regions effectively suppress infections. For example, the total effective reproduction number is reduced by 17%, 25%, and 36% if the infection rate is suppressed to 10% in the top-5, -10, and -20 highest-risk 1 km-squares in period (a), respectively. With regard to real restrictions, the 1st SoE successfully reduced the population density in the downtown region. The SoEs reduced the exponents; however, the change amount of the exponent before and after SoEs also decreased.

**Figure 5.**
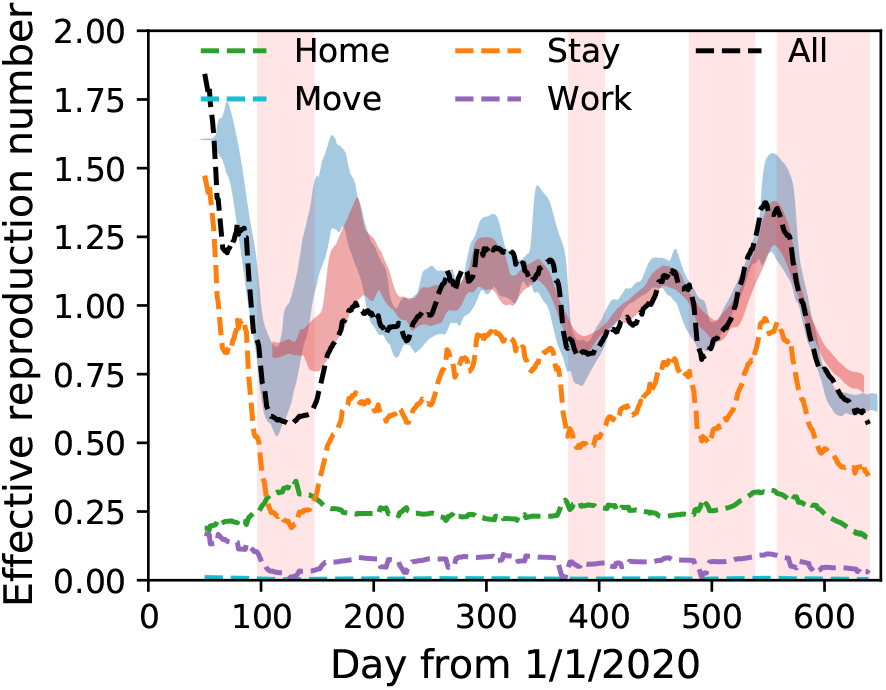
Components of effective reproduction number in the model. Empirical data are indicated by blue and red bands for all infections and severe cases, respectively. The stay-out activity gives the largest component in most time series.

**Figure 6.**
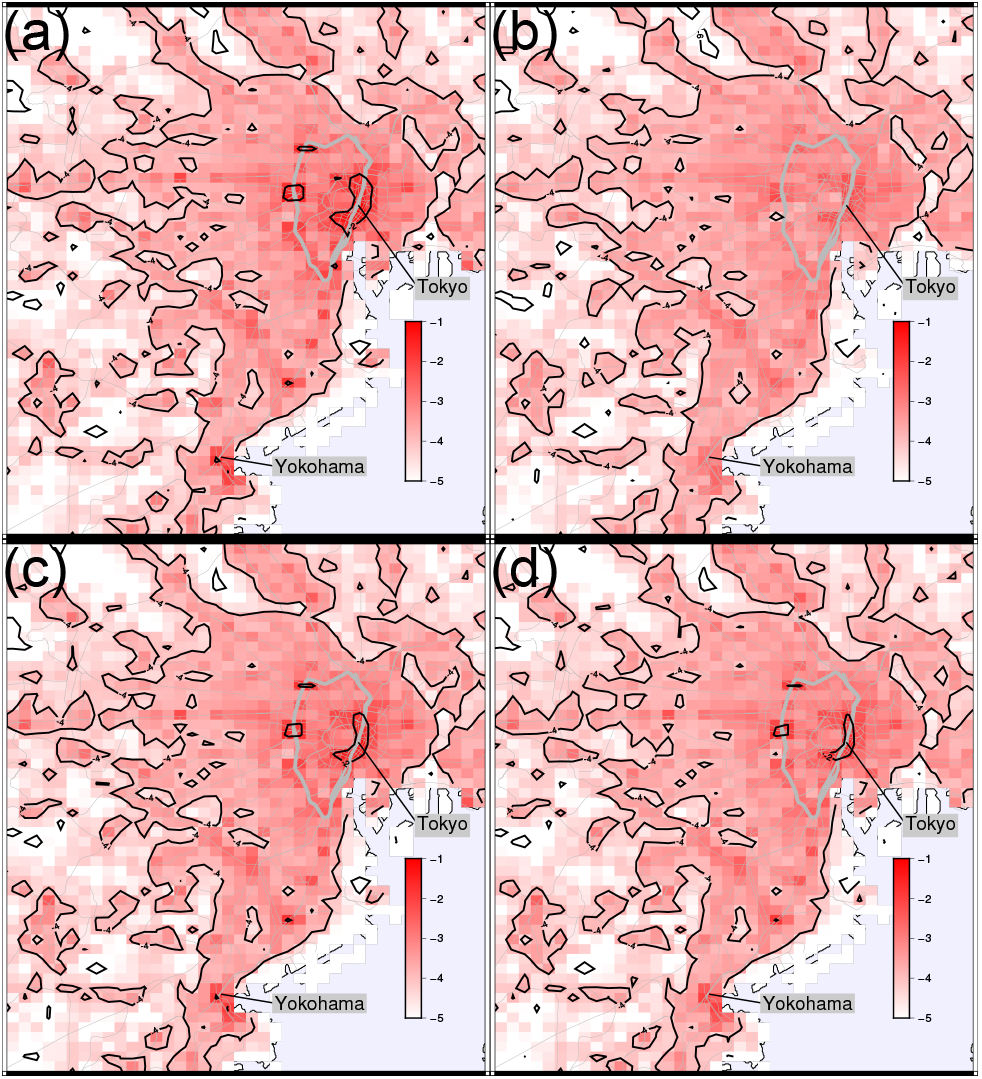
Effective reproduction number map of Tokyo metropolitan area (a) two months before 1st SoE, (b) during 1st SoE, (c) two months before 2nd SoE, and (d) during 2nd SoE. The Yamanote line is indicated by a bold grey closed curve, and other railways are indicated by fine grey curves. The colour represents the values on a log-10 scale. The sum of all regions is the overall effective reproduction number. High-risk zones are concentrated around Tokyo station and the Yamanote line. The maps are generated by Generic Mapping Tools (GMT) 6.0.0^49^ (https://www.generic-mapping-tools.org/).

**Figure 7.**
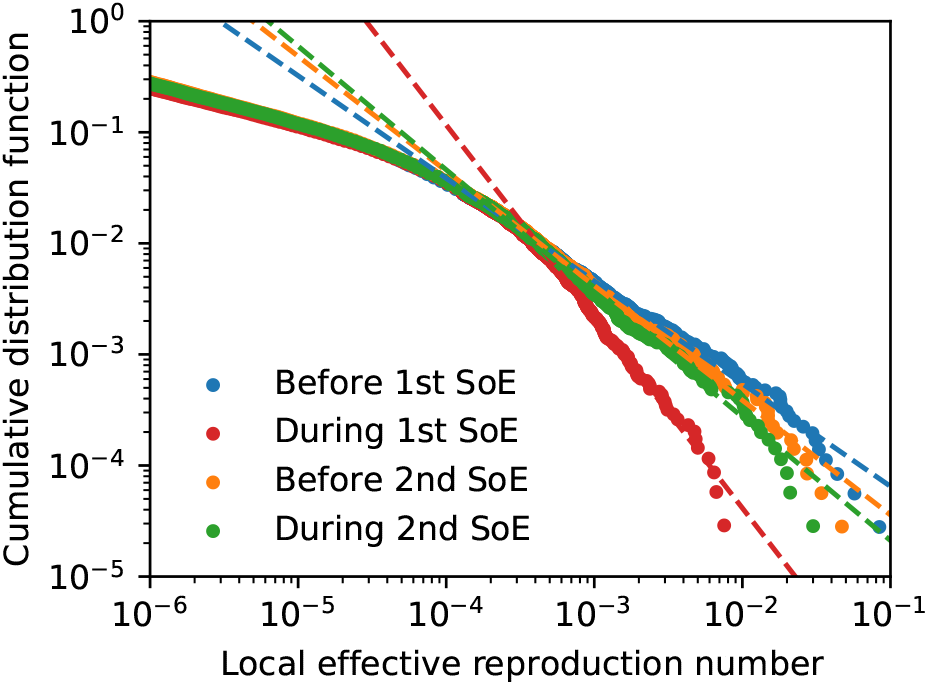
Cumulative distribution function of effective reproduction number at each location on a log-log scale (a) two months before 1st SoE, (b) during 1st SoE, (c) two months before 2nd SoE, and (d) during 2nd SoE. Power-law fitting functions are plotted as dotted lines. The exponents are (a) -0.92, (b) -1.72, (c) -1.03, and (d) -1.11. The sum of the distribution is the total effective reproduction number.

The effective reproduction number for each infected person^47,48^ is defined by the mean number of people to whom they would spread the infection; the average for all people gives the overall effective reproduction number. We calculate it using the time series of the activity and the location in a day by the following relation:

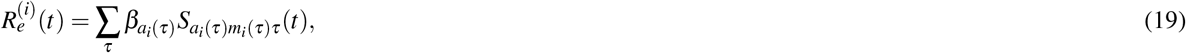

where *τ* is the time label with 15 min intervals, and *a*_*i*_(*τ*) and *m*_*i*_(*τ*) represent the history of person *i*. We refer to this value as the GPS-based individual effective reproduction number. Fig. 8 shows the cumulative distribution function (CDF) of 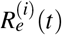, where we collect all data for (a) two months just before the 1st SoE, (b) during the 1st SoE, (c) two months just before the 2nd SoE, and (d) during the 2nd SoE. The distribution is approximated by truncated power laws. The exponents between -1 and 0 in (a) and (c) clearly indicate that significant cluster infections or superspreaders dominate the total infection. In fact, the top 10% of people contribute to (a) 58%, (b) 38%, (c) 54%, and (d) 50% of the overall effective reproduction number. A comparison before and during SoEs reveals that the cut-off does not change while the exponent is decreased. This implies that the effects of the SoE are represented by the reduction of the exponent in the CDF power law; it results in the suppression of the overall effective reproduction number. The cut-off corresponds to the largest infection clusters or superspreaders, which was not suddenly changed by SoEs; however, we observe that it decreased slowly from the 1st to the 2nd SoE.

**Figure 8.**
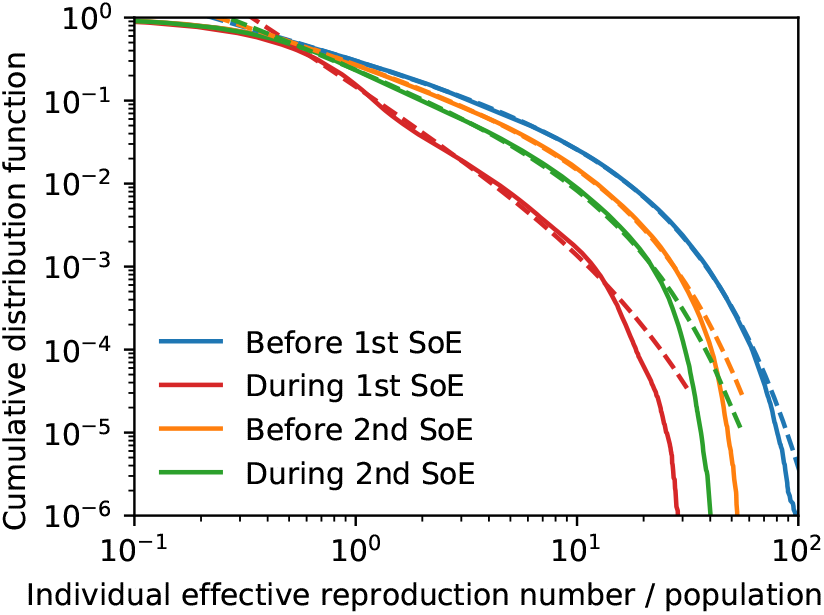
Cumulative distribution function of individual effective reproduction number on a log-log scale (a) two months before 1st SoE, (b) during 1st SoE, (c) two months before 2nd SoE, and (d) during 2nd SoE. Fitting functions are plotted as dotted lines: (a) 0.33*r*^−0.76^ exp(−0.078*r*), (b) 0.16*r*^−1.72^ exp(−0.080*r*), (c) 0.30*r*^−0.86^ exp(−0.105*r*), and (d) 0.26*r*^−1.03^ exp(−0.108*r*), where 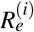 is denoted by *r*. The mean of the distribution is the effective reproduction number.

## Discussion

In this study, we verified human-activity-dependent COVID-19 infection rates using smartphone GPS data. We classified human activity patterns into four types, ‘home’, ‘move’, ‘stay-out’, and ‘work’, and estimated the number of social contacts for each daily activity in the Tokyo metropolitan area. Then, we derived an equation from the classical SIR model for GPS data with activity information to be used directly. The model successfully predicted effective reproduction numbers for future reporting. We demonstrated that infection risk is the highest when the people are not at home or work or not moving. By quantitatively understanding the effect of human mobility on infection spread, we distinguished the impact of the Delta variant and vaccination. Furthermore, we derived formulas that divide the effective reproduction number into the contributions from each location or individual. These formulas enabled us to observe the distributions of infection risk. As applications, we present an effective reproduction number map and GPS-based individual effective reproduction numbers, whose distributions obey the power law or truncated power law.

The model provides a comprehensive understanding of infection spread of epidemics. A previous research by T. Yabe et al.^9^ investigated the COVID-19 spread in the Tokyo metropolitan area during 2020 in detail and discovered the nonlinear relation between the effective reproduction number and the contact index, the number of social contacts among people not in their homes. The nonlinear relation is explained by the change in the ratio of the population moment of stay-out to that of work and move. Another previous research conducted by S. Rüdiger et al.^15^ shows that the non-uniformity in the infective contact network has an important role as well as the total number of social contacts. The difference from this study in approach is the origin of the non-uniformity in social contacts: the non-uniform distribution of the infectious people in their study (c.f. it can break the assumption (A3)), and the type of the social contacts in this study.

We validated the model in the following two ways: demonstrating the stability of prediction by using a shorter fitting span and comparing the results with a previous study. We have already shown the result of fitting in the span 200 ≤ *t <* 500, but checking the result in another shorter span is meaningful as cross-validation. Here we fit the parameters in the shorter span

200 ≤ *t <* 300 to check the model stability as shown in Supplementary note 4. We observe that both errors in the parameters and prediction are reasonable despite the data containing non-stationarity. Another comparison is with the airborne transmission dynamics^27^ in possible situations in daily life. Previous research by M. Prentiss et al.^27^ indicated that the infection risk is increased by 10 times or more depending on the use of masks, filtration, and talking. Their results are consistent with ours that stay-out activity, including eating at restaurants, enhances the infection rate more than 10 times.

The assumptions (A1)-(A4) determines the limitation of our model, but relaxing some of them can lead to a new application or complex model for epidemics. If we do not adopt the assumption (A1), the effective reproduction number is decreased to the ratio (1 −*R*(*t*)*/N*). This is because the possibility of infection in each location decreases to the same ratio. For the assumption (A3), the non-uniformity in the infective contact network is considered. Although it is impossible to track each person across the days in the GPS dataset for privacy protection, preferences of people with high infection risk could be assumed or observed in another dataset to estimate the spatial distribution of infected people.

For future research, the infection spread to the whole country could be simulated. In the second wave of the pandemic in Japan, after the infection spread among metropolitan areas, it exploded into countryside areas^28^. We have already started the research of modelling the infection spread across distant regions. The model will be adopted in other metropolitan areas to validate the model further, and the parameters possibly dependent on the regions will be determined and discussed. After analysing the bulk property of epidemics in each area, we will introduce the interaction between the regions into the model, using the GPS data of travelers.

For practical applications, the result of this study may be helpful for evidence-based policy making for epidemic control, such as non-pharmaceutical interventions under pandemics. The following questions will be answered: “How does the human mobility decrease in target areas affect the effective reproduction number?” and “What intervention is the most effective under given constraints?”. In our estimation, the contribution from the downtown area fills a large amount of the effective reproduction number. It will be possible to suggest the locations where activities such as stay-out should be limited enough to realize a goal effective reproduction number. This model can also be applied to real-time forecasting of the infection spread and assessing the individual infection risk. We are preparing a web page that will illustrate the infection risk of each location and time on a map, using the epidemiological data and the semi-real-time GPS data provided by Agoop. People can check the infection risks of visiting places on the web page. Also, we suggest implementation in a smartphone application that tracks the user’s location each time. The smartphone application downloads the estimated infection risks of the visiting places each day. After the day, the individual effective reproduction number, which represents both the risk of transmitting infection and being transmitted, is given a notice to the users for risk management. Finally, we are proceeding with a study to estimate the trade-off relations between the infection spread and the economy, using economic data on each site together. We will analyse the relationship between human mobility and the companies’ sales in the leisure industry, such as restaurants, department stores, and traveling. The sales of those companies that are generated in each location are estimated as a function of human mobility, with the data of the spatial distributions and the mean expenditure of such restaurants or stores. This investigation bridges the effective reproduction number with the economy in each location.

## Data Availability

The data that support the findings of this study are available from Agoop Corp. but restrictions apply to the availability of these data, which were used under license for the current study, and so are not publicly available. Data are however available from the authors upon reasonable request and with permission of Agoop Corp.

## Acknowledgements

We thank Kenta Yamada, Yukie Sano, Takashi Shimada, and Takahiro Nishi for the helpful discussions. We thank Agoop for providing the GPS datasets. This work was supported by the Tokyo Tech World Research Hub Initiative (WRHI) Program of the Institute of Innovative Research, Tokyo Institute of Technology.

## Data availability

The data that support the findings of this study are available from Agoop Corp.^30,50^ but restrictions apply to the availability of these data, which were used under license for the current study, and so are not publicly available. Data are however available from the authors upon reasonable request and with permission of Agoop Corp.^30,50^

## Author contributions

M.T. was the project leader and directed the entire research plan and writing of the manuscript. Y.S. pre-processed the raw GPS data and appended the activity information. J.O. analysed the pre-processed GPS data, developed the models, performed the numerical calculations, and wrote the manuscript. H.T. improved the data analysis and modelling methods and revised the manuscript.

