## Supplemental Notes for "Direct modelling from GPS data reveals daily-activity-dependency of effective reproduction number in COVID-19 pandemic"

### Supplementary Material

Jun'ichi Ozaki, Yohei Shida, Hideki Takayasu, Misako Takayasu

**Supplementary note 1 | Estimation of population moment using 1km-square data.**

We consider the case of using the 100m-squares instead of the 1km-squares because the social contact is still inhomogeneous within the 1km-squares. We estimate the population moment $M_a^{100\mathrm{m}}(t)$, defined by the 100m-square data of GPS, from the 1km-square data because the GPS data does not have enough resolution in home squares for privacy protection and enough user number for the data limitation. Here we think of 100m-squares labeled by $m_{100\mathrm{m}}$ within a 1km-square marked by $m_{1\mathrm{km}}$, and a partial sum $\sum_{m_{100\mathrm{m}} \in m_{1\mathrm{km}}} \left(S_{am_{100\mathrm{m}}\tau}(t)\right)^2$. We approximate it as a function of the population of 1km-square $S_{am_{1\mathrm{km}}\tau}(t)$. In Fig. S1, for all 1km-square in Japan, we plot the partial sum of the population moment $\sum_{m_{100\mathrm{m}} \in m_{1\mathrm{km}}} \left(S_{m_{100\mathrm{m}}\tau}(t)\right)^2$ conditioned by the 1km population $S_{m_{1\mathrm{km}}\tau}(t)$, where activities do not condition them, and the home activity is removed because of its low resolution. The figure implies that if we focus on high-density areas, the partial sum is approximated by

$$\sum_{m_{100\mathrm{m}} \in m_{1\mathrm{km}}} \left(S_{am_{100\mathrm{m}}\tau}(t)\right)^2 \simeq 0.024 \left(S_{am_{1\mathrm{km}}\tau}(t)\right)^2, \tag{1}$$

where

$$\sum_{m_{100\mathrm{m}} \in m_{1\mathrm{km}}} S_{am_{100\mathrm{m}}\tau}(t) = S_{am_{1\mathrm{km}}\tau}(t). \tag{2}$$

If the population is homogeneous in the 1km-square, the coefficient should be 0.01, not 0.024. The population inhomogeneity in 1km-squares makes the coefficients 2.4 times larger. The population moment is calculated as

$$M_a^{100\mathrm{m}}(t) = \gamma^{-1} \sum_{m_{1\mathrm{km}} \in A, \tau \in t} \left[ \sum_{m_{100\mathrm{m}} \in m_{1\mathrm{km}}} \left(S_{am_{100\mathrm{m}}\tau}(t)\right)^2 \right] \simeq 0.024\gamma^{-1} \sum_{m_{1\mathrm{km}} \in A, \tau \in t} \left(S_{am_{1\mathrm{km}}\tau}(t)\right)^2 = 0.024 M_a(t). \tag{3}$$

Therefore the infection rate $\beta_a^{100\mathrm{m}}$ using the 100m-squares is calculated as

$$\beta_a^{100\mathrm{m}} = \beta_a / 0.024, \tag{4}$$

where $\beta_a$ is the infection rate in the 1km-square case.

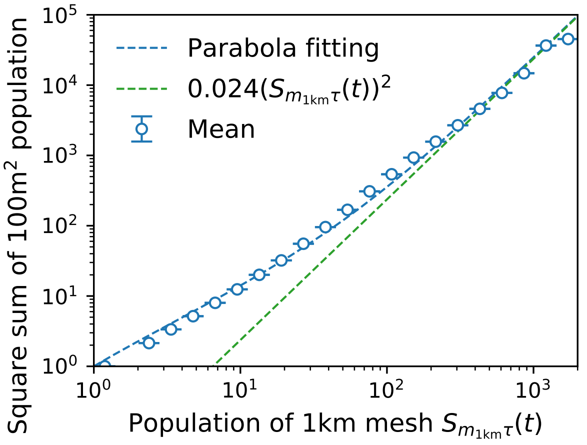


**Supplementary Figure S1.** Partial sum of the population moment conditioned by the 1km population without the home activity in Japan. The error bars show the standard deviation of the mean.

##### Supplementary note 2 | GPS data example.

We show an example of the GPS data used in this study. The GPS data is provided as csv data. The used or important record list is: "*dailyid, year, month, day, dayofweek, hour, minute, latitude, longitude, os, accuracy, speed, course, prefcode, citycode, mesh100mid, home_prefcode, home_citycode, workplace_prefcode, workplace_citycode*". An example of our GPS data is: "4c6e13d9a51b6091, 2020, 4, 15, 3, 14, 30, 35.671145, 139.748757, Android, 600, 0, 90, 13, 13101, 5339450959, 13, 13213, 13, 13103". Details of the data header are described in Table S1.

The data provider estimated the users' home and work city using the GPS tracking data over several days, which is not included in the provided data. The provided data were fully anonymized. All user IDs were renewed every midnight, so any tracking of individual persons across the days is impossible. For data intervals, 94% of the data is taken within under 15 minutes (1 January 2020).

**Supplementary Table S1.** GPS data columns that are used or important. The data is provided as csv files.

| header | description |
|---|---|
| *dailyid* | the daily ID of the user changed everyday |
| *year, month, day, dayofweek, hour, minute* | time, *dayofweek* is 1 (Mon.) to 7 (Sun.) |
| *latitude, longitude* | the location of the user |
| *os* | the operating system of the smartphone |
| *accuracy* | the position acculacy (m) |
| *speed* | the estimated velocity (absolute value) (m/s) |
| *course* | the estimated direction of the velocity (0-360) |
| *prefcode, citycode* | the prefecture and city code of the present location |
| *mesh100mid* | 100m mesh (100m square or grid) label defined by the Japanese Ministry of Land, Infrastructure, Transport and Tourism |
| *home_prefcode, home_citycode* | the estimated prefecture and city of the homeplace |
| *workplace_prefcode, workplace_citycode* | the estimated prefecture and city of the workplace |

**Supplementary note 3 | Spatial data coverage map.**

Here, we compare population density maps of the GPS data and census data. Figures S2 and S3 show the population of the GPS and census data around Tokyo and Figs. S4 and S5 plot the population of the whole Tokyo metropolitan area. The color intensity is proportional to the population density of the residents, where the GPS data of population is observed at 5 a.m. on 1st April 2020, and the census data of population used the national census of Japan in 2015 (provided by https://www.e-stat.go.jp/en/). The GPS data is renormalized to fit the whole population in each prefecture. The density patterns are consistent, meaning that the population observed in the GPS data represents the actual population well. Still, the population from the GPS has more contrast, which implies that the smartphone users (who also use the applications provided by Agoop Corp.) are concentrated in the high-density population areas, although these two data have a five-year difference.

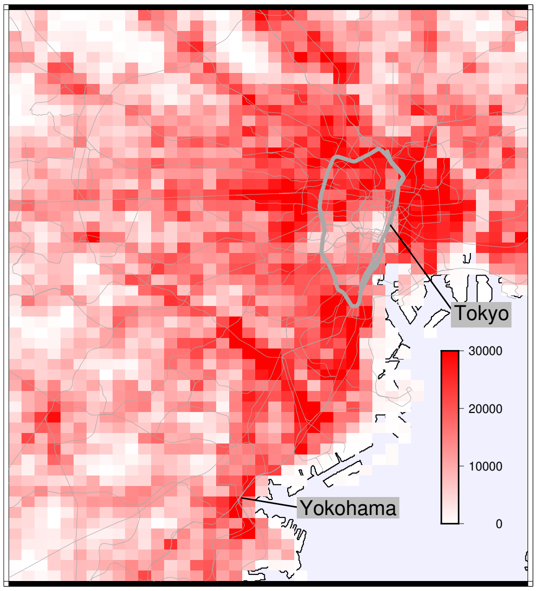


**Supplementary Figure S2.** The population density of the residents around Tokyo from the GPS data at 5 a.m. on 1st April 2020.

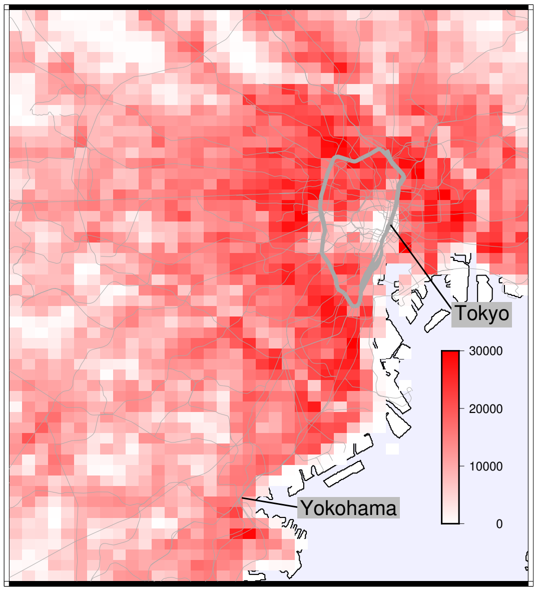


**Supplementary Figure S3.** The population density of the residents around Tokyo from the national census of Japan in 2015.

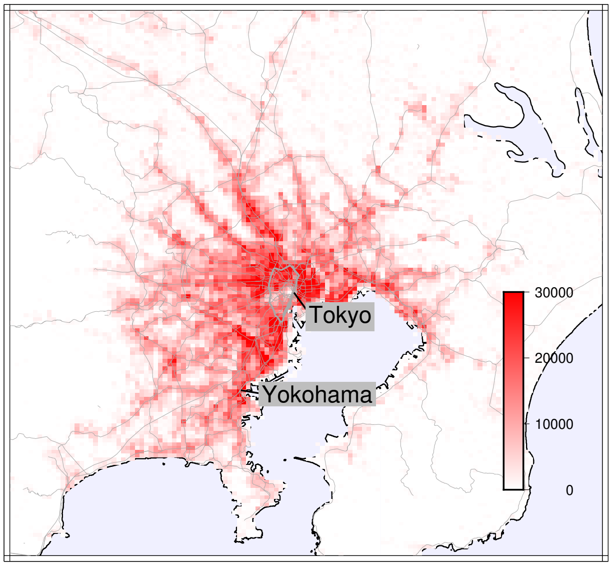


**Supplementary Figure S4.** The population density of the residents in the whole Tokyo metropolitan area from the GPS data at 5 a.m. on 1st April 2020.

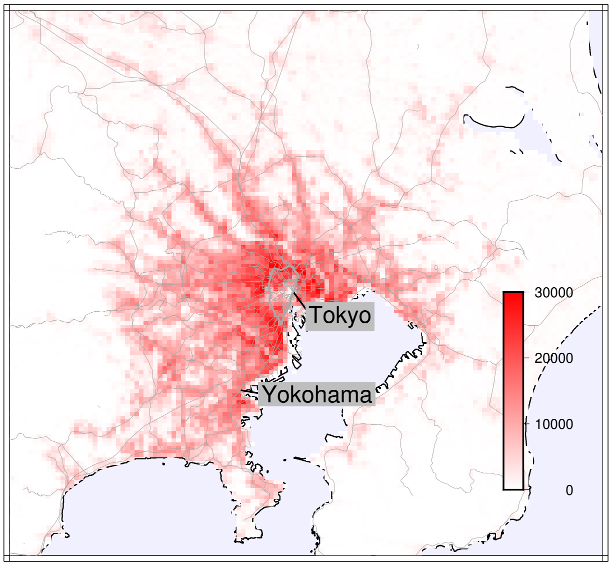


**Supplementary Figure S5.** The population density of the residents in the whole Tokyo metropolitan area from the national census of Japan in 2015.

**Supplementary note 4 | Validation for the method and model.**
We validate the method for the effective reproduction number. This study calculates the effective reproduction number as the ratio of the number of newly confirmed cases

$$R_e^{\text{data}}(t) = \frac{I^{\text{new}}(t+\gamma^{-1})}{I^{\text{new}}(t)}, \tag{5}$$

where we take a 7-day moving average for the number of newly confirmed cases $I^{\text{new}}(t)$ (Cislaghi method) [*Alessandro, A. & Tommi, Tech. Rep. JRC121343, Publications Office of the European Union (2020).* ]. To validate the method in our data, we compare the result with the Cori method [*Cori, A., Ferguson, N. M., Fraser, C. & Cauchemez, S., Am. J. Epidemiol. 178, 1505–1512*]. Figure S6 shows the comparison. The timeseries of the Cori method are ahead by 2 days, but both are consistent. Most of the timeseries of the Cislaghi method well approximate that of the Cori method, although the peaks are sharper in the Cislaghi method.

Next, we discuss the parameter fitting in the model. In the main result of this study, we have fit the data to determine the infection rate parameters $\beta_a$ during the timespan $200 \leq t < 500$. For the validation, we do it using the part of the timespan $200 \leq t < 300$ ($\Delta T = 14$ is not changed). The parameters are $\beta_{\text{home}} = (1.7 \pm 0.3) \times 10^{-7}$, $\beta_{\text{move}} = \beta_{\text{work}} = (1.2 \pm 0.5) \times 10^{-7}$, and $\beta_{\text{stay}} = (2.5 \pm 0.4) \times 10^{-6}$. These parameters differ from those of the span $200 \leq t < 500$ by up to 2.5 standard deviations, but this error is reasonable because the data contains non-stationarity. A predicted result using these parameters is shown in Fig. S7, which is consistent with the main result of $200 \leq t < 500$. Here, the effects of the Delta variant and the vaccination is not included.

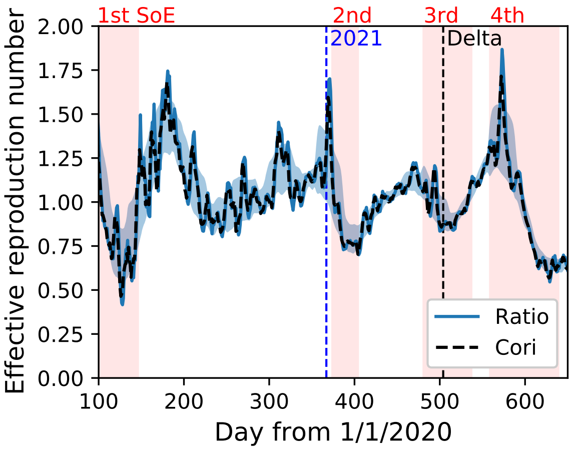


**Supplementary Figure S6.** Comparison between the method used in this study (Cislaghi method) and the Cori method. 'Ratio' indicates the method in this study, and 'Cori' does the Cori method.

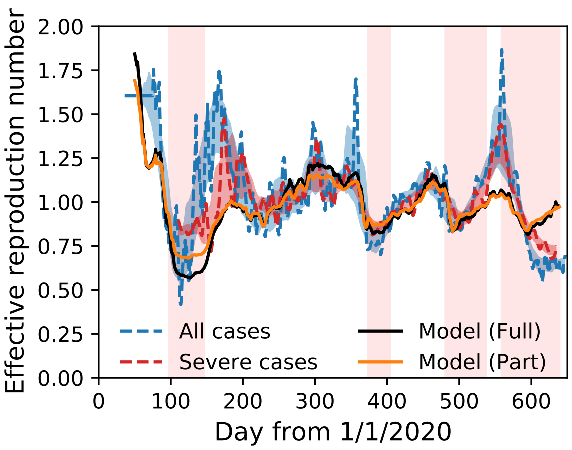


**Supplementary Figure S7.** Effective reproduction number of all infections and severe cases in the Tokyo metropolitan area compared to the model results of fitting during the full span (full, $200 \leq t < 500$) and the partial span (part, $200 \leq t < 300$).
